# Head and Body Pose Classification for Understanding Sleep Behaviour in People Living with Dementia using Video and a Novel Multi-Head Attention-Driven Deep Learning Architecture

**DOI:** 10.64898/2026.04.29.26351379

**Authors:** Sarmad Al-Gawwam, Marta M Pineda, Kiran K G Ravindran, Ciro della Monica, Giuseppe Atzori, Ramin Nilforooshan, Hana Hassanin, Victoria Revell, Derk-Jan Dijk, Kevin Wells

## Abstract

Sleep posture is known to be relevant to various sleep disorders, such as sleep apnea, but is not often quantified in sleep monitoring systems. We address this with a novel vision-based approach, which is robust to the challenging conditions (variable lighting, partial occlusions, variable geometry) of in-bed monitoring. This paper proposes a novel, attention-driven deep learning framework for the robust classification of head and body pose from infrared (IR) video streams during sleep of older people and people living with Alzheimer’s. Our architecture integrates a pre-trained convolutional backbone with a novel Multi-Head Channel-Spatial Attention (MH-CSA) module. The MH-CSA mechanism hierarchically identifies salient features by first capturing multi-scale spatial context using parallel heads with varied dilation rates, and then adaptively recalibrating feature importance via integrated Squeeze-and-Excitation blocks. To specifically address class imbalance, the model is optimized using a Dynamic Class-Balanced Focal Loss, which forces the network to focus on hard-to-classify examples from underrepresented classes. Whilst most prior sleep analysis work is developed using data from healthy younger participants, our system was developed and validated on a nocturnal sleep dataset of older adults and people living with Alzheimer’s disease, with IR video synchronized to clinical video-Polysomnography (vPSG). For head position classification, the system achieved an F1-score of 91% for older adults and 90% for people living with Alzheimer’s; for body pose prediction, the scores were 91% and 89% for the respective cohorts. These results demonstrate significant potential for application in understanding sleep behavior and informing appropriate sleep interventions.

## I. INTRODUCTION

**S**LEEP is an essential part of our daily lives and has a significant impact on our health and quality of life. Sleep disturbances are highly prevalent in people who developed neurodegenerative diseases such as Alzheimer’s and Parkinson’s disease. Thus, these abnormalities present as an early biomarker in the pre-clinical phase, preceding the onset of cognitive decline and are very common neuropsychiatric symptoms accompanying neurological disorders [1, 2].

A high frequency of body movements during sleep is characteristic of disrupted sleep, which is associated with poor self-reported sleep quality [3]. Sleep disorders, such as sleep disordered breathing (SDB), cause a wide range of symptoms such as pauses in breathing accompanied by hypoxia, awakenings, excessive daytime sleepiness and cognitive impairment. Such disorders can also be exacerbated by certain body poses during sleep that impact the severity of the disorder [4], for example, several studies have shown a close relationship between supine head/body pose, and the severity of sleep apnea [5, 6]. Moreover, it has been demonstrated that people with neurodegenerative disease spend more time in the supine head position than people with normal cognition [7].

Beyond global body posture, head posture during sleep has been shown to substantially modulate Obstructive Sleep Apnea (OSA) severity, with lateral rotation of the head while the body remains supine significantly reducing the apnea–hypopnea index compared to a fully supine head-and-body position; this demonstrates that head position exerts an independent influence on upper-airway collapsibility in positional OSA [8]. In a large population-based cohort, time spent in the supine sleeping position was positively associated with higher AHI, and this positional effect on OSA severity was disproportionately greater in overweight individuals than in normal-weight or underweight participants, underscoring the interaction between sleep posture and obesity [9]. Quantifying and characterizing movements are also important to correctly classify and distinguish parasomnias, such as Rapid eye movement (REM) sleep Behavior Disorder, from sleep-related movement disorders [10].

Objective sleep monitoring, including head and body pose, is crucial for the identification and diagnosis of sleep disorders, including sleep apnea, which may require targeted clinical attention. The gold standard AASM method to record and diagnose sleep disorders is polysomnography (PSG), where data are collected by attaching an array of sensors and electrodes to the participant’s body. The sleep recordings are then scored by highly trained polysomnographic technologists. Although PSG offers detailed physiological data, it is intrusive, expensive, and time-consuming to set up and score as it requires laboratory visits, specialized wearable sensors and professional installation. Furthermore, PSG can impact the sleep of the participant under investigation because of unfamiliarity with the new environment [11]. These characteristics prevent PSG from being used for the longitudinal study of sleep behavior. Therefore, the primary motivation for this work is to enable a non-contact methodology that can be deployed in the familiar home environment and that can successfully characterize sleep and body and head position in particular in sensitive populations (such as those living with dementia) who may not tolerate wearable devices.

There has been significant prior work in non-contact sensing modalities to address the in-bed pose estimation problem. These include approaches based on pressure mapping systems [12, 13], depth sensing [14], and infrared imaging [15–17]. Recent advances in machine vision, particularly in the domain of human pose estimation, have presented significant opportunities for developing novel, non-contact monitoring systems [18]. However, the direct application of these computer vision techniques to in-bed pose estimation is constrained by the challenging environmental conditions, including variable illumination (up to and including darkness) and persistent occlusions from bedding, which degrade the performance of state-of-the-art human pose estimation algorithms in these scenarios [19].

To overcome these issues, we introduce a non-contact monitoring solution for use within an non-contact framework. Our proposed system utilizes an infrared (IR) camera combined with an AI Head and Body pose classification model described here. This model employs a novel deep-learning architecture that combines a deep convolutional backbone with a hierarchical attention mechanism for robust and accurate head and body pose classification from IR images. The main contribution of this work can be summarized in three-parts:

- We propose a novel ResNet-Multi-Head attention architecture applied to body and head recognition during realistic sleep conditions with a low computational complexity attention mechanism. This AI model is robust to body and head position occlusion by blankets, variable illumination, and camera viewpoint.
- To improve the localization of discriminative features of body and head, we propose a Multi-Head Convolutional Attention mechanism which employs a shared depth-wise convolution across parallel attention heads to capture discriminative features of different body and head positions efficiently.
- To address the severe class imbalance inherent in sleep poses during model training, we introduce a Dynamic Class-Balanced Focal Loss. This loss function integrates two key principles: a class-balancing weighting scheme based on the effective number of samples in different batches, and a dynamic modulating factor that adaptively adjusts the focal loss for each class based on model confidence. This dual mechanism allows the model to shift its focus onto minority classes during model training and hard-to-classify examples dynamically.

To evaluate the performance of the model, we utilize overnight video data of sleep behavior drawn from cognitively healthy older people and a group of adults living with Alzheimer’s disease and ground truth labels derived from consensus-based expert annotation. This is in contrast to prior work [13, 20, 21], which is often based on data acquired from healthy, younger individuals; whereas we deliberately sought to capture the disturbed sleep characteristics of older cohorts.

## II. RELATED WORK

Recent research in sleep monitoring has demonstrated the effectiveness of non-contact-based systems in capturing and analyzing sleep-related data such as heart rate (HR), respiration rate (RR), movement, and posture [20, 22]. For instance, Zhu et al. [22] proposed a privacy-protected defocused infrared camera system for real-time sleep parameter measurement that extracts HR and RR via spatially-redundant remote photoplethysmography (rPPG, non-contact optical vital sign estimation from skin color changes), motion signals for respiration and body activity, and classifies five sleep postures with ResNet-18, achieving strong correlations (*R* ≈ 0.91 for HR, *R* ≈ 0.97 for RR) and 94.5% posture accuracy even under bed sheet occlusions. Tam et al. [20] further advanced blanket-robust posture classification with SaccpaNet, a separable atrous convolution-based cascade pyramid attention network pretrained on Red–Green–Blue (RGB) depth data from 150 participants across four blanket thicknesses classes. The application of deep learning, and specifically attention mechanisms, has been explored in various biomedical and healthcare contexts. Vaswani et al. [23] introduced the transformer architecture, which relies on multi-head selfattention to model complex dependencies in sequential data. Ahmed et al. [24] developed an IoT-based smart healthcare system using Mask-RCNN for noninvasive patient discomfort detection from top-view IP camera RGB video. The system extracts 17 body keypoints, transforms them into six major body parts via data mining association rules, achieving 94% true-positive rate with 7% false-positive rate. Liu et al. [13] introduced the Simultaneously-Collected Multimodal Lying Pose (SLP) dataset with RGB, Long Wave-length Infrared (LWIR), depth, and pressure map modalities for in-bed pose monitoring under adverse conditions (darkness, full occlusion), enabling state-of-the-art 2D body pose estimation. However, the study settings in these methods are constrained to well-illuminated environments and to cases with little to no occlusions. We have previously explored various methods to detect and analyse body pose, i.e., [25]. Mahvash et al. [25] were the first to demonstrate that a ResNet combined with transfer learning can successfully detect pose of a body obscured by a bed covering. The method was found to be more accurate than polysomnography belt-based body pose detection. However, recent further experimentation demonstrated that this approach was unable to successfully detect head poses with similar levels of performance. This motivated the development of a more complex model architecture to study variations of head pose and body pose in older adults and those living with dementia who are particularly susceptible to sleep apnea. We developed and tested our model using gold-standard, ground truth data collected in cognitively healthy older people and people living with Alzheimer’s (PLWA) with varying degrees of sleep apnea.

## III. Data Acquisition

Before participant recruitment and acquisition of data from the cognitively healthy group, this study was submitted for ethical review to the University of Surrey Ethics Committee and received a favourable ethical opinion (UEC 2019 065 FHMS, 2 August 2019). For the study involving PLWA we sought prior ethical review from the UK NHS Health Research Authority (HRA) and were granted a a Favorable Ethical Opinion, (22/LO/0694, 2nd November 2022). All participants provided written informed consent before participation.

Details of the demographic and sleep characteristics of the two sets of participants can be found in Table I. Prior to sleep recordings, cognitively healthy older participants reported a range of co-morbidities and other long standing health issues (such as osteoarthritis, hypertension etc). However, none of them were considered to have mental impairment (which may impact sleep behaviour) as measured by the mini-mental state examination for clinically significant cognitive impairment (all with MMSE scores *>*23). The PLWA had a diagnosis of prodromal or mild Alzheimer’s Disease based on clinical history, cognitive tests, and CT/MRI imaging. PLWA comorbidities included hypertension, osteoarthritis, asthma and type II diabetes. Further details can be found in [26]. Two datasets containing one full night video-PSG recording per participant were used in the development and testing of the proposed AI model: (i) recordings from 35 cognitively healthy older adults and (ii) recordings from 6 PLWA. In both datasets, participants underwent 10 hours of video-PSG recorded using the Somnomedics SOMNO-HD device and the Somnomedics (IR) (Somnomedics Gmbh, Germany) Home Sleep Camera.

**TABLE I:** Demographic and sleep characteristics of participants across dataset splits. Note: CI: Cognitively healthy; PLWA: People living with Alzheimer’s disease; BMI: Body mass index; AHI: Apnea-hypopnea index; TST: Total sleep time; SE: Sleep efficiency (100 X (TST/Recording time)); WASO: Wake After Sleep Onset; SOL: Sleep Onset Latency.

| Characteristic | CI Train/Valid | CI Test | PLWA Test |
| --- | --- | --- | --- |
| No. of participants | 31 | 4 | 6 |
| Sex (M/F) | 19/12 | 2/2 | 3/3 |
| Age (years, Mean ± SD) | 70.6 ± 4.9 | 73.0 ± 3.2 | 71.3 ± 4.7 |
| BMI (kg/m<sup>2</sup>, Mean ± SD) | 27.0 ± 4.9 | 25.0 ± 3.4 | 26.8 ± 8.6 |
| AHI (events/h, Mean ± SD) | 20.3 ± 16.0 | 17.1 ± 12.3 | 22.5 ± 19.5 |
| TST (Minutes, Mean ± SD) | 389 ± 65.20 | 362.3 ± 74.72 | 299 ± 6.5 |
| SE (% , Mean ± SD) | 71.22 ± 10.79 | 69.1 ± 7.73 | 61.5 ± 13.8 |
| SOL (Minutes, Mean ± SD) | 14.6 ± 11.7 | 19.3 ± 20.4 | 33 ± 43 |
| WASO (Minutes, Mean ± SD) | 137.95 ± 54.88 | 138.75 ± 19.68 | 151.3 ± 49.26 |

Individual sound attenuated temperature-controlled single sleep rooms were used for sleep recordings of the first 18 cognitively healthy participants. For the remaining 17 cognitively healthy participants and the second test set of 6 PLWA, sleep recordings were acquired in our Living Lab facilities within the Surrey Sleep Research Centre which combines a domestic home environment with unobtrusive instrumentation including IR video monitoring.

The first set of 35 cognitively healthy participants was used to train, validate and initially test the model. The second dataset of 6 PLWA was used as a second holdout test set for the model to further explore its generalizability to a different cohort acquired at a different time point. Further details on the split and training regime appear in Section IV.

PSG sleep summary estimates were computed for the lights off to lights on interval as per AASM guidelines [27]. The extended period in bed, which results in more wake after sleep onset, provided a strong basis to test the system for monitoring movements and poses in both wake and sleep. Sleep stages (wake [W], rapid eye movement (REM), stage N1 of non-REM sleep [N1], stage N2 of non-REM sleep [N2], and stage N3 of non-REM sleep [N3]) were scored at 30s intervals using the DOMINO software environment by two independent scorers from which a consensus hypnogram was generated. IR video recording was captured with a Somnomedics active IR camera system (Somnomedics Home Sleep Camera, Somnomedics Gmbh, Germany) at 25 frames per second (fps), with a video resolution of 800 × 600 over the recording period for each participant. The camera position and orientation in each of the recording bedrooms was varied and created a different view of the bed for each participant, but typically 2.2m height was used, offset sideways by around 1m or positioned directly over the center of the bed. These video recordings form the basis of the core data used in this work. Fig.1 presents examples of each of the pre-defined head and body poses from different participants. Body and head positions for all participants were annotated using individual video frames down-sampled from the original 25 frames per second video stream to one frame per 30s interval. For body pose labels, left or right body labels were assigned to a frame when the inferred roll angle between the shoulders and the bed towards the participant’s left or right side, exceeded 45 degrees. If the estimated angle was less than this, then the pose was deemed to be supine. Body yaw and pitch angles were ignored. A similar heuristic rule was used to determine the head position, but in this case, nose position was used instead of the shoulders.

**Fig. 1:**
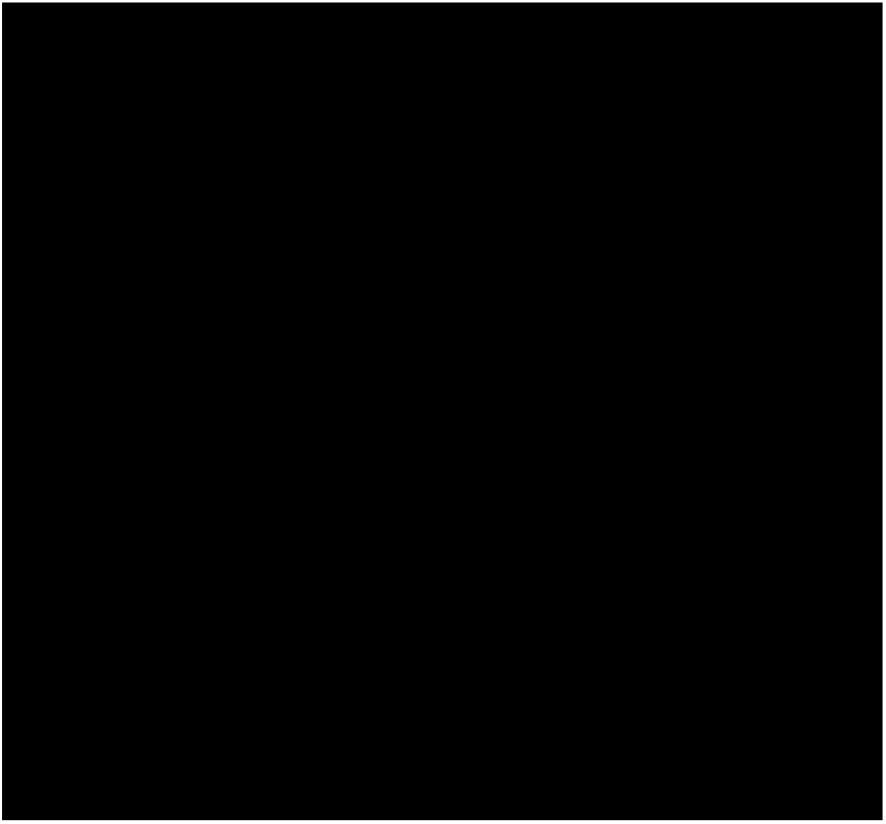
Examples of body and head poses in three participants: (A) supine, (B) right, (C) left. Note that on some occasions, the blanket partially or fully obscures the head and/or body. Figure redacted due to MedRViV submission rules. Access to the detail of this figure is available from the corresponding author.

Following random allocation to training, validation and test partitions, consensus labels from 2 expert observers (1 of which was a Registered Polysomnographic Technologist) were generated for all test cases and 9 of the training cases. This produced agreement scores spanning 86-100%. Differences were then arbitrated to arrive at a consensus agreement of 100% for the labelled cases.

## IV. PROPOSED METHOD

Fig. 2 shows a schematic diagram of the proposed method summarized below. We start with IR data, which are downsampled to one frame per 30-second epoch. The down-sampled 2D IR video camera data are transformed using homography to a common viewpoint to standardize the head and body orientations across different individuals and to remain robust to different camera deployments. Then, a pre-trained ResNet-50 model [28] is leveraged as a head/body image feature extractor (FE) to extract complex appearance information from the input video frames. To enhance the model’s perception of task-relevant features under challenging conditions, such as partial occlusion and low-light, we introduce a novel Multi-Head Channel-Spatial Attention (MH-CSA) module, as shown in Fig. 3. Specifically, our proposed MH-CSA module computes multi-scale spatial importance maps of different head and body positions using parallel heads with varying dilation rates, allowing the model to capture context from different receptive fields. Furthermore, we integrate a Squeeze-and-Excitation (SE) block [29] within each attention head. This creates a hierarchical attention mechanism that first identifies where to focus spatially, that is, it highlights the most relevant pixel regions corresponding to the head or body within the image frame. This then recalibrates those feature channels that are most important within that identified region. Further details on the internal architecture are illustrated in Fig. 3. Finally, the dynamic focal loss is used during model training to dynamically address the class imbalance problem. Further details appear in sections A and B. Finally, to mitigate the negative impact of class imbalance inherent in clinical sleep data during model training, we employ a custom Dynamic Class-Balanced Focal Loss. This function adaptively re-weights the contribution of each sample, allowing the model to focus on hard-to-classify examples from minority classes.

**Fig. 2:**
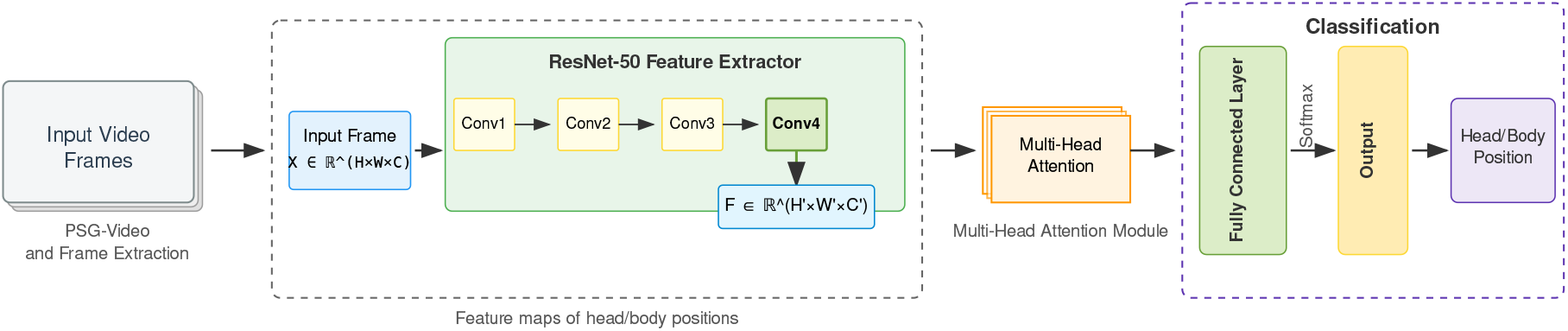
Schematic diagram of the proposed methodology. A single-channel input IR frame (*X* ∈ ℝ^*H×W ×C*^) is first transformed using homography. To ensure compatibility with the ResNet-50 backbone [28], which expects a 3-channel input, this single channel is replicated to create a 3-channel tensor (*X* ∈ ℝ^*H×W ×*3^). The pre-trained ResNet-50 then acts as a feature extractor, producing a high-dimensional feature map (*F* ∈ ℝ^*H*^*′*^*×W*^ *′*^*×C*^*′*). This feature map is subsequently processed by the Multi-Head Channel-Spatial Attention (MH-CSA) module to generate the final pose classification.

**Fig. 3:**
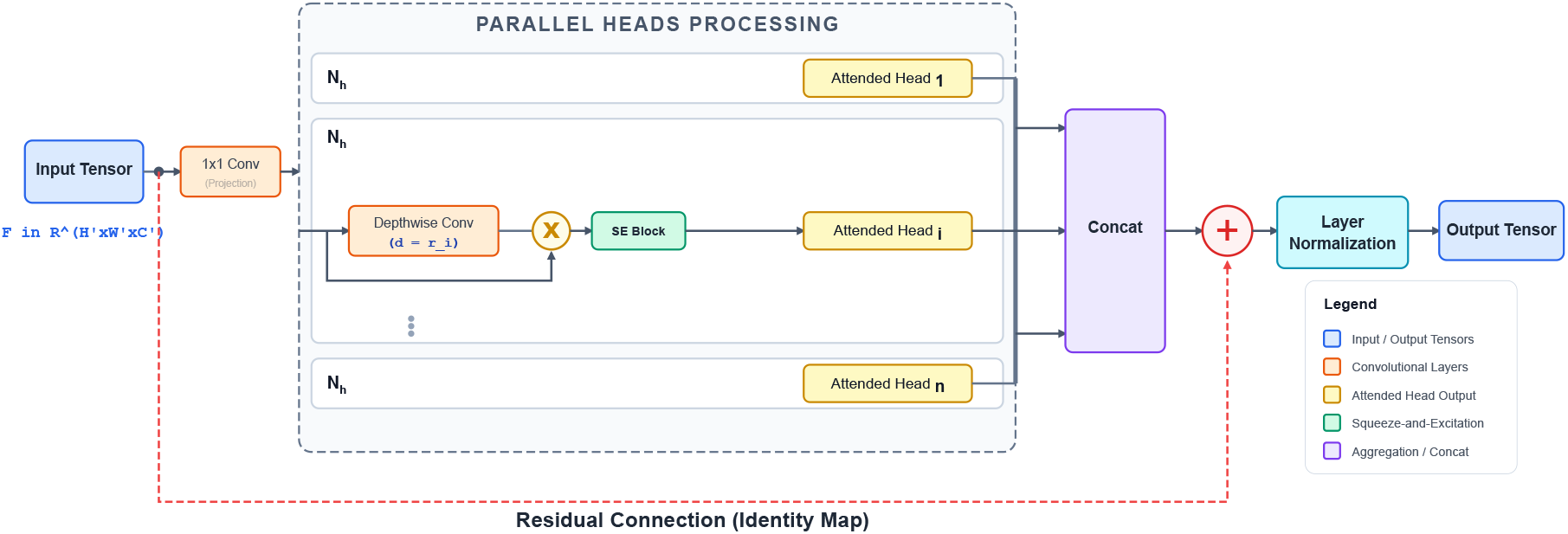
Detailed architecture of the proposed attention block. The input tensor is processed by multiple heads in parallel. Within each head i, the input is used in two ways: (a) it is passed through a Conv2d layer and a dilated depthwise convolution to generate a spatial gate, and (b) a corresponding group of channels is sliced to form the feature group. The gate is applied to the features via element-wise multiplication. The outputs are then concatenated and passed through a residual connection, enabling multi-scale context aggregation with low computational overhead.

### A. Head/Body Image Feature Extractor Module

The starting point of our proposed architecture is a deep convolutional feature extractor (Fig.2). To this end, we employ a transfer learning technique, utilizing the pre-trained ResNet-50 model as the feature extractor. The ResNet backbone is pre-trained on the ImageNet dataset, comprising millions of common objects and scenes. A significant domain gap exists between these source images and the target clinical sleep domain. Consequently, while the early layers of the network learn generalizable, low-level feature detectors (e.g., for edges, textures, and gradients) that are highly transferable, the deeper layers learn features specialized to the original ImageNet classes, limiting their direct utility for our sleep pose classification task. Therefore, we utilize the output of the fourth convolutional stage (Conv4) of the ResNet-50 backbone as the primary feature extractor layer. This intermediate stage provides semantically rich, high-level features that are sufficiently general to be adapted for robust head and body pose classification. The input can be formally described as a tensor *X* ∈ ℝ^*H×W ×C*^, where *H* and *W* represent the height and width of the image in pixels, respectively, and *C* = 3 is the number of channels. Where monochrome IR data are used then only a single channel may be needed. But as the ResNet architecture requires 3 channel RGB data, C is kept for consistency for this and other applications.

The feature extractor then produces a high-dimensional feature map *F* ∈ ℝ^*H*^′^×*W*^ ′^×*C*^′, where the primed dimensions indicate a change dimensions after the convolutional layers of the ResNet-50 network. This tensor, which encapsulates a rich hierarchy of learned visual patterns, serves as the direct input to the subsequent Multi-Head Channel-Spatial Attention (MH-CSA) module, which is tasked with identifying and amplifying the most salient features for the pose classification task.

### B. Multi-Head Channel-Spatial Attention (MH-CSA)

Transformer-based models have demonstrated remarkable success, primarily due to the Multi-Head Self-Attention (MHSA) mechanism’s ability to capture important information from data [23]. In the conventional MHSA, the intermediate feature map *F* ∈ ℝ^*H*^*′*^*×W*^ *′*^*×C*^*′* is flattened spatially and treated as a sequence of *N* = *H*^*′*^ × *W* ^*′*^ tokens. These tokens are projected into Query (*Q*), Key (*K*), and Value (*V*) representations to compute dot-product attention:

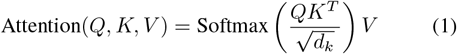

where *d*_*k*_ is the dimension of the Key vectors, used to prevent the dot products from growing too large and to stabilize gradients during training. The primary limitation of this mechanism is its computational and memory complexity. The dot-product attention in Equation 1 must compute a pairwise similarity score between all *N* tokens, resulting in an attention matrix of size *N* × *N*. This operation has a computational and memory complexity that scales quadratically (i.e., *O*(*N* ^2^)) with the number of tokens *N* = *H*^*′*^ × *W* ^*′*^. This quadratic growth with respect to the input’s spatial resolution makes MHSA computationally expensive for many applications that require real-time processing of high-resolution sensor data. Inspired by this challenge, the Multi-Head Channel-Spatial Attention (MH-CSA) block was designed (see Fig. 3). The proposed MH-CSA block directly addresses this limitation by replacing the quadratic-complexity dot-product operation with a highly efficient, linear-complexity convolutional approach, designed to provide enhanced performance for head and body position recognition.

#### 1) MH-CSA Architecture

The MH-CSA block processes an input feature map through four main stages: a) initial feature projection, b) parallel multi-head hybrid attention, c) feature aggregation and normalized residual connection.

##### a) Feature Projection

Given an input tensor *F*, we first apply a point-wise (1 × 1) convolution to project it into a richer feature space, enhancing its representative capacity. This can be formulated as:

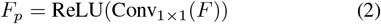

where *F*_*p*_ is the projected feature map.

##### b) Multi-Head Hybrid Attention

The core innovation of the proposed MH-CSA lies in its parallel attention heads, which perform a hybrid of spatial and channel attention without incurring the cost of standard self-attention. The projected feature map *F*_*p*_ is split along its channel dimension into *N*_*h*_ parallel attention heads. For each head *i* ∈ {1, …, *N*_*h*_ }, a feature slice 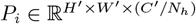 is processed. Here, *C*^*′*^*/N*_*h*_ denotes that the total channels *C*^*′*^ are divided equally among the *N*_*h*_ heads. This process is detailed as follows:

First, for each head a *multi-scale spatial attention map* is generated. Instead of using *Q* and *K* matrices, we employ a computationally efficient *DepthwiseConv2D* layer with scaled dilation rate *d*_*i*_. This allows each head to capture contextual information at a different scale, followed by a sigmoid activation function. This process can be expressed as:

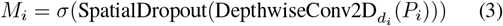

where *M*_*i*_ is the spatial attention map for head *i*, and *σ* is the sigmoid function. The set of dilation rates 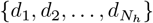 allows the block to concurrently model fine-grained local details and broader, long-range spatial relationships.

Next, these attention weights are multiplied element-wise by the same input channel slice. This gating mechanism adaptively emphasizes important spatial locations:

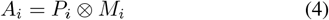

where ⊗ denotes element-wise multiplication.

Finally, to further enhance feature representation, a *Squeeze-and-Excitation (SE) block* [29] performs dynamic channelwise feature recalibration on the spatially-attended features *A*_*i*_. The SE block determines the importance of each channel within the head and adaptively re-weights them, allowing the model to focus on the most informative features. The output of the head is thus:

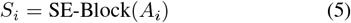

##### c) Feature Aggregation and Normalization

The outputs *S*_*i*_ from all *N*_*h*_ heads are concatenated along the channel dimension. This aggregated feature map is then combined with the input feature map *F* (the input to this block) via a residual connection and stabilized with Layer Normalization to produce the final output *F*_out_.

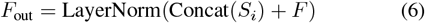

where Concat(S_*i*_) denotes concatenation over all heads S_*i*_ ∈ {1, …, *N*_*h*_ }.

#### 2) Dynamic Class-Balanced Loss

The distribution of head and body positions in the studied dataset presents two challenges for deep learning models. First, the dataset exhibits a natural class imbalance, where certain poses occur more frequently than others. Standard loss functions, such as Categorical Cross-Entropy (CCE), can become biased towards these majority classes, leading to poor performance on minority classes, which may be clinically significant. Second, as training progresses, the model is dominated by the loss from a large number of “easy” examples (i.e., clear head or pose without obstructions or edge cases), which can overwhelm the learning signal from the few “hard” examples that show more challenging head and body positions, which are critical for improving model robustness.

To address these challenges, this paper proposes a novel loss function called Dynamic Class-Balanced Loss (DCBL) that dynamically balances the loss contribution during model training. DCBL results in more stable training behaviors and efficiently classifies the majority and minority classes, making it an ideal strategy for imbalanced datasets.

The core objective is to re-weight the standard Cross-Entropy loss on a per-sample basis. The final loss for a given sample is defined as:

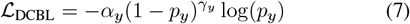

where *y* denotes the ground-truth class label, *p*_*y*_ is the model’s predicted probability for the ground truth class, the two key modulating factors, *α*_*y*_ and *γ*_*y*_, are designed to handle class imbalance and hard examples, respectively.

First, the Class-Balancing term, *α*_*t*_, assigns a higher weight to samples from minority classes. It is calculated based on the effective number of samples for each class, which is a smooth measure of class frequency [30]. This ensures that the model is penalized more for misclassifying rare samples, forcing it to learn their features more effectively. Second, the Focal Loss term, 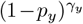, addresses the issue of easy versus hard examples. For well-classified examples where the model is confident (*p*_*y*_ → 1), the 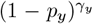 factor becomes very small, down-weighting the loss contribution from these easy samples. Conversely, for hard-to-classify examples (*p*_*y*_ → 0), this factor approaches 1, preserving the loss and focusing the training on these challenging cases.

The key novelty in our implementation is making the focusing parameter, *γ*_*y*_, dynamic. Instead of using a fixed *γ* as a hyperparameter, it is adjusted for each class based on an exponential moving average of the model’s prediction probabilities for that class. This allows the loss function to automatically increase the focus (i.e., use a higher *γ*) on classes that the model finds more difficult over time. By integrating these components, our loss function creates a robust training objective.

### C. Dataset and Preprocessing

The data processing pipeline was designed to extract frames corresponding to the center of each 30-second epoch of the PSG recording. To create a representative image for each epoch, we extracted the middle video frame of each 30-second sleep epoch to reduce computational cost while preserving essential postural information. This method ensures a consistent capture of the patient’s posture within each time window. To focus the analysis on sleep-specific head and body positions, all epochs classified by the PSG as ‘Wake’ were excluded from the final dataset. To reduce noise and reduce lighting effects while maintaining essential structures of body and head positions, Contrast Limited Adaptive Histogram Equalization (CLAHE) was applied to the extracted frames to heighten image contrast without losing important image information. The extracted frames were then resized to a uniform resolution of 224 × 224 pixels.

### D. Experimental Evaluation

All models in this study were implemented using the TensorFlow framework, run on an NVIDIA RTX 4080 GPU. The model was trained using overnight IR video of participants (Table I) drawn from the cognitively healthy older adults cohort using the AdamW optimizer [31] with *β*_1_ = 0.9 and *β*_2_ = 0.999. We employed a batch size of 32. To ensure stable training and promote convergence, we utilized a combined learning rate scheduling strategy consisting of a linear warmup phase for the initial epochs, followed by a Cosine Annealing decay schedule. The learning rate *η*_*e*_ at any given epoch *e* is defined as:

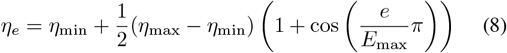

where *η*_max_ was set to an initial learning rate of 1 × 10^−4^, *η*_min_ is the minimum learning rate, and *E*_max_ is the total number of epochs designated for the decay schedule. An early stopping mechanism, monitoring the validation loss with a patience of ten epochs, was implemented to prevent overfitting and save the best-performing model. Validation used three participants from this same dataset, and an initial hold-out test set of four participants was used to evaluate the model for both head and body poses using consensus pose labels, also drawn from the cognitively healthy group. The hold-out test was selected in order to represent a range of sleep apnea cases, ranging from AHI of 7.6 to 35.1 (Table I). This was evaluated using metrics derived from the resulting confusion matrices for the Head and the Body pose analysis.

To evaluate the generalizability of the proposed architecture, a second hold-out test set representing video of overnight sleep drawn from six PLWA was also used on the aforementioned model (with the same weights frozen after training as used in the initial test set). The use of data collected in a different acquisition environment and participants with a different neurological profile (expected to produce different sleep patterns from those in the first cohort) was used to examine the generalizability of the model. The classification performance for both head and body positions was evaluated using metrics derived from the resulting confusion matrices. We also compared the performance of the model predictions for body position with the output from the integrated body position sensor (Somnomedics SOMNO HD, Somnomedics GmbH, Germany), used routinely within PSG to estimate body pose. Finally, we compared, for the first time, differences in body pose vs head pose in older adults.

### E. Evaluation Metrics

The performance of the proposed model was quantitatively evaluated using the following metrics:

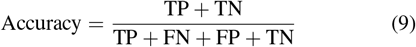

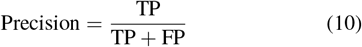

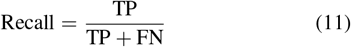

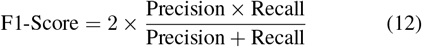

where TP, TN, FP, and FN represent the total number of true positive predictions, true negative predictions, false positive predictions, and false negative predictions, respectively.

### F. Computational complexity

To assess the suitability of the proposed architecture for applications where computational efficiency is as critical as predictive accuracy, a thorough performance analysis was conducted. All training and inference benchmarks were executed on a single NVIDIA RTX 4080 GPU. The proposed model, including the ResNet-50 backbone and the MH-CSA module, requires approximately 8.24 GFLOPs (Giga Floating Point Operations) for a single forward pass on a 224 × 224 × 3 input tensor. It is critical to note that the novel MH-CSA module is designed for efficiency; through the use of point-wise and depthwise convolutions, it adds minimal computational overhead relative to the ResNet-50 backbone, making it a powerful and lightweight addition. The model was trained for 25 epochs, with an average training time of approximately 4 minutes per epoch. For real-time application viability, the model’s inference speed was benchmarked. With a batch size of one, the model achieved an inference speed of 14.27 frames per second (FPS), corresponding to an average processing time of 70.09 ms per frame. The model’s inference performance is therefore considered suitable for deployment, as the processing time per frame (70.09 ms) is suitable compared to the 30-second epoch interval used for real-time monitoring in a clinical or home setting.

## V. EXPERIMENTAL RESULTS

### A. Distribution of Ground Truth Head and Body Positions for the Two Test Cohorts

The total number of ground truth body pose counts was substantial for both Test Set 1 and Test set 2 (Table II). The supine position was more frequent in the PLWA than in the CI cohort (60.5% vs 40.5%). The left body position was the least frequent in both cohorts. For head position a similar pattern was present, although the supine head position was less frequent than the supine body position. Thus overall, PLWA spent more time in the supine position.

**TABLE II:**
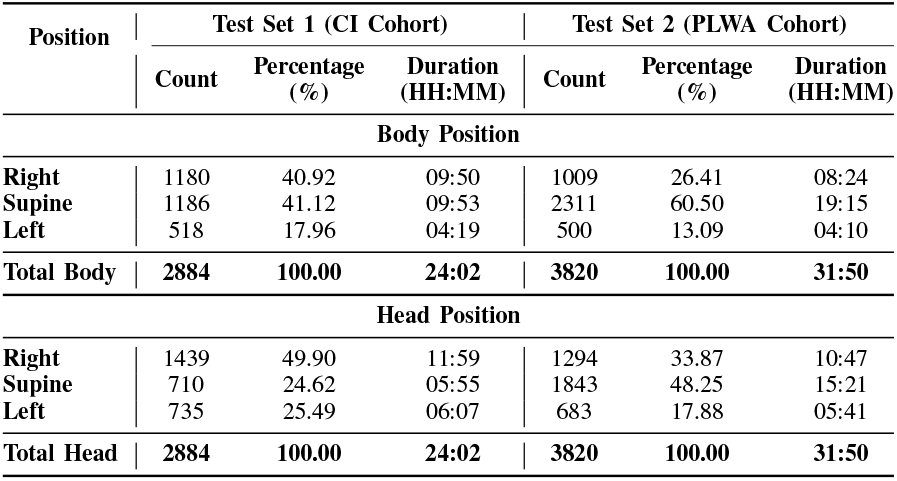
Distribution of Ground Truth Head and Body Positions for the Two Test Cohorts. The data reflects the total number of frames (Count), the percentage distribution within each cohort, and the total time spent in each position during sleep.

### B. Head and Body Model Performance

In Fig. 4, we present an example of AI head model predictions against ground truth. In the same figure we also present body model pose predictions against the consensus ground truth (as described in Section III) and those labels obtained using the to infer body position from the SomnoMedics PSG system for one of the cognitively healthy test participants. The performance of our Head Model tested on cognitively healthy individuals and PLWA test sets is presented in Fig. 5a and Fig. 5b, respectively. From this, a variety of metrics for the Head Model can be calculated, as shown in Table III. The Body Model was also tested on both cognitively healthy individuals and PLWA, similarly to the Head model. The resulting confusion matrices are presented in Fig. 5c and Fig. 5d, and the derived metrics are reported in Table III.

**TABLE III:** The performance of Head and Body models across the test sets of head and body positions. Numbers represent the mean performance across all participants. Note: CI: Cognitively Healthy, PLWA: People Living With Alzheimer’s.

| Metric | Head Model |  | Body Model |  |
| --- | --- | --- | --- | --- |
|  | CI Test Set | PLWA Test Set | CI Test Set | PLWA Test Set |
| Sensitivity (Recall) | 92% | 90% | 91% | 92% |
| Specificity | 96% | 95% | 95% | 96% |
| F1 Score | 91% | 90% | 91% | 89% |
| Precision | 91% | 90% | 91% | 87% |

**Fig. 4:**
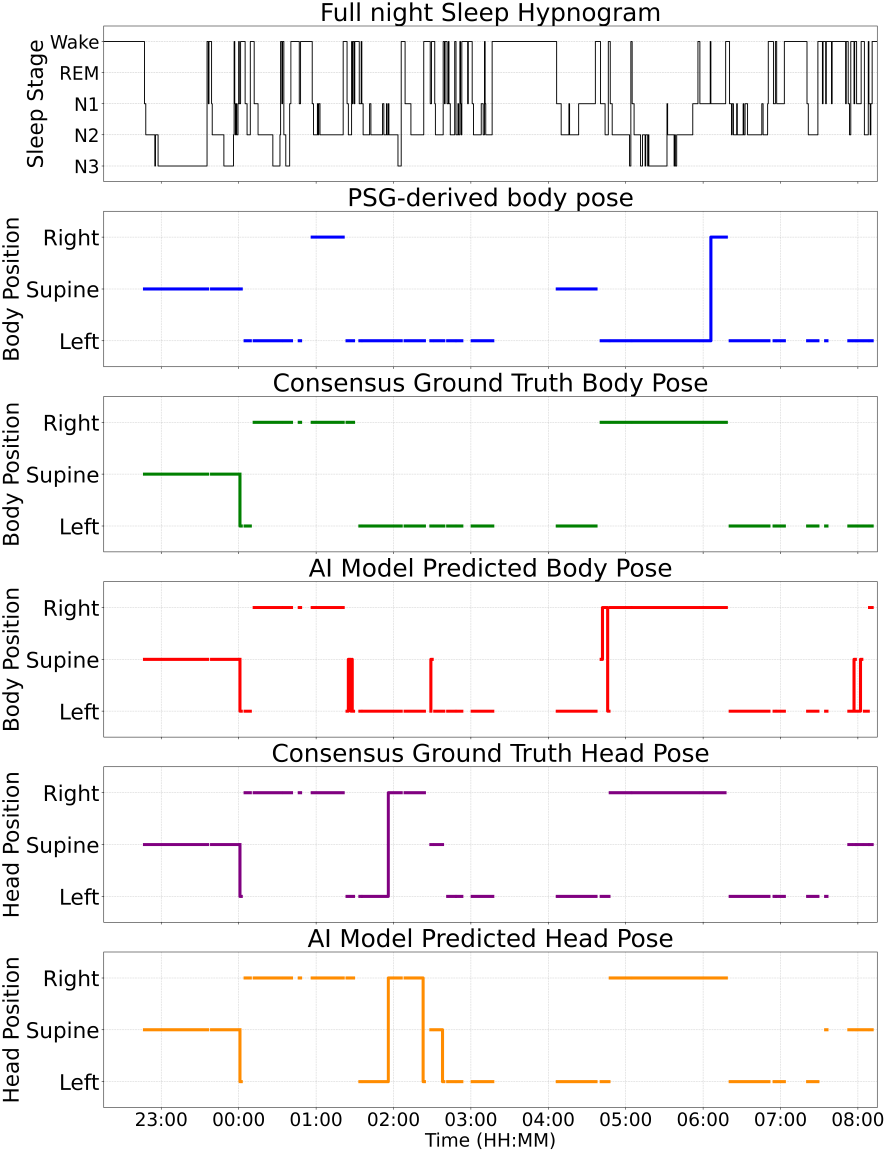
Illustration of model performance and PSG-derived hypnogram during sleep states. From top: (i) PSG-derived hypnogram, (ii) body pose labels derived from the PSG system’s body position sensor, (iii) consensus ground truth labels for body pose, (iv) AI model predicted body pose predictions, (v) consensus ground truth labels for head pose, (vi) AI model predicted head pose predictions. Note that body and head pose were not quantified during wake.

**Fig. 5:**
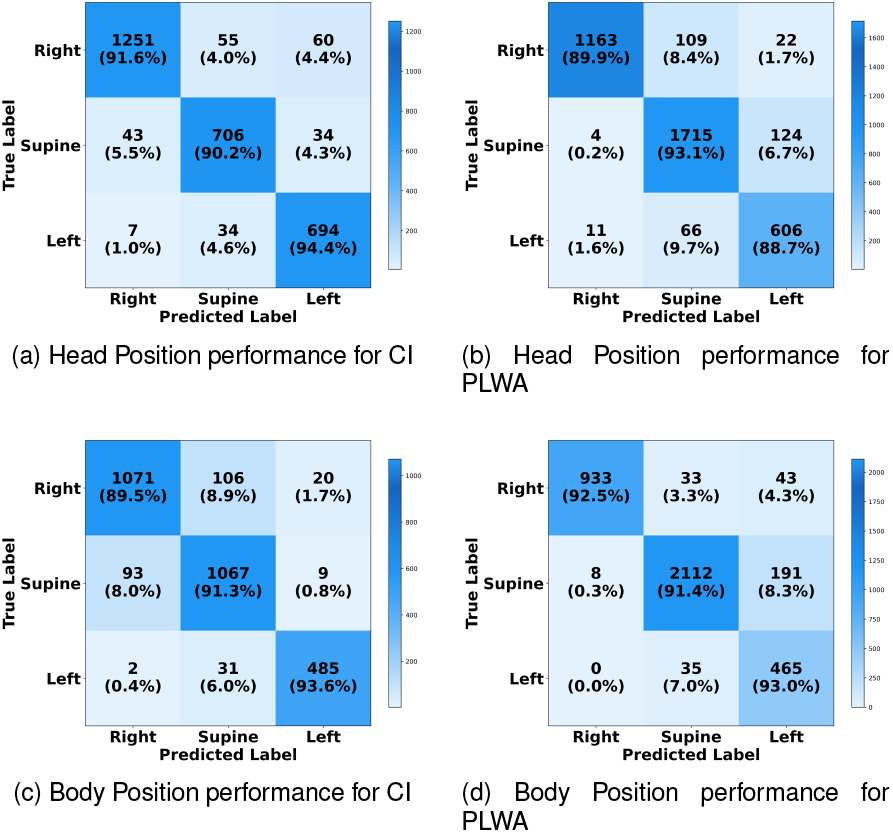
Confusion matrix of the proposed Body and Head model. Note: CI: Cognitively Healthy, PLWA: People Living With Alzheimer’s.

In Fig. 6 we compare the AI model accuracy against the PSG system in determining body position for all participants (cognitively healthy and PLWA), benchmarked using the previously described manual labels. This clearly shows the higher accuracy of the AI model in the vast majority of cases. We found that for some of the participants, the AI model accuracy was only ∼ 80%. Visual inspection of the frames revealed that, these participants had covered their whole body, which may explain this level of performance for those cases.

**Fig. 6:**
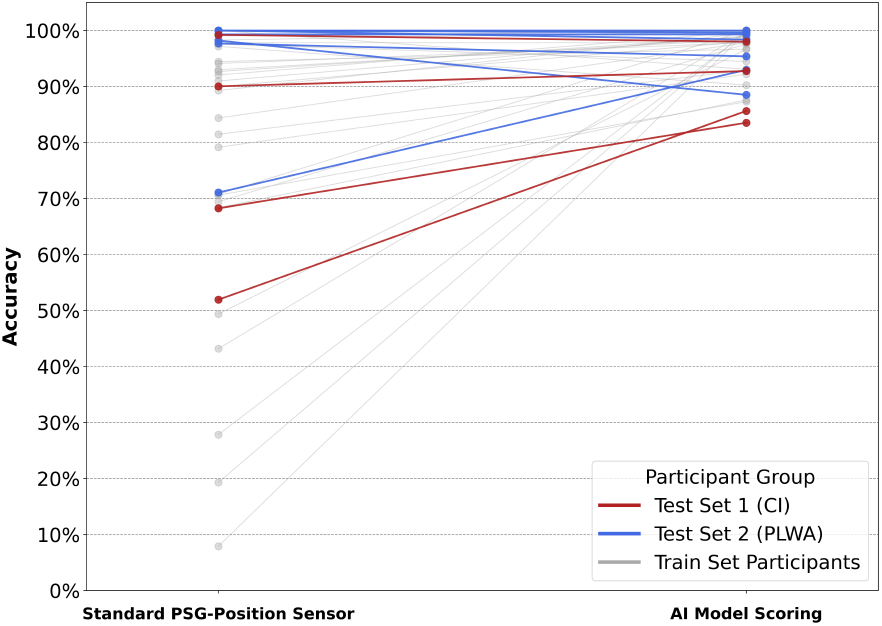
Body model pose accuracy for cognitively healthy particpants and PLWA compared to accuracy of the standard PSG position sensor for all participants.

### C. Performance of MH-CSA module on Head and Body Classification

To validate the effectiveness of the proposed Multi-Head Channel-Spatial Attention (MH-CSA) module, an ablation study was conducted. To better illustrate the role of the MH-CSA in head position classification, the t-distributed neighbor embedding (t-SNE) method was employed to visualize the outputs from various layers. Fig. 7 displays these visualization results, with test data being randomly subsampled to enhance the clarity of the presentation. Initially, the input data were randomly distributed, and as a result, the data cannot be easily separated into visually distinct clusters (see Fig.7 A). After applying MH-CSA layer, left, right and supine head positions were effectively distinguished (see Fig.7 B). The differentiation among positions became more pronounced following the attention layer. Overall, the separability of the head positions during sleep improved progressively after the application of the MH-CSA layer in the network.

**Fig. 7:**
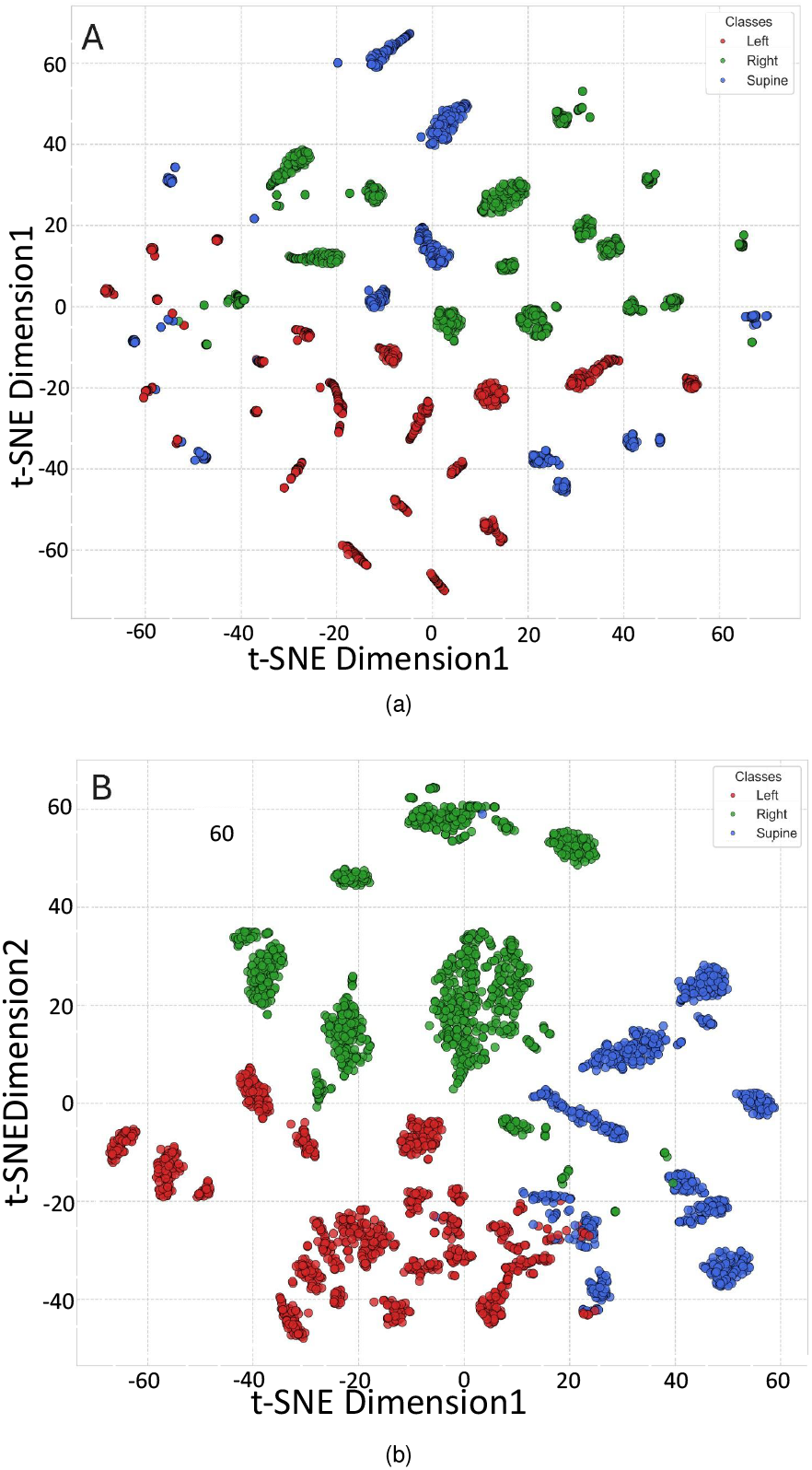
Visualization of the separability of outputs using t-SNE. Panel A: Input features. Panel B: Multi-Head attention module output.

Table IV presents a detailed performance comparison between the proposed model architecture and a baseline model that uses Resnet-50 as a feature extractor and discards the MH-CSA module. The results show that the integration of the MH-CSA module yields a substantial and consistent improvement across all metrics. This indicates that the attention mechanism enhances the model’s ability to focus on relevant features within the IR images, strengthening classification performance. For the Head Model, the impact is particularly significant in the PLWA test set. In this case the Precision increases dramatically from 66.0% to 90.0%. This suggests that the MHCSA module is highly effective at minimizing false positive predictions. Furthermore, Sensitivity shows an increase from 61.0% to 90.0%, demonstrating that the model is more effective at detecting true positive cases when attention is applied, which is vital in ensuring that clinically significant postures are not misclassified. These individual metric improvements culminate in a significant F1-score uplift. For the Head Model, the F1-score on the PLWA test set significantly improves from 63.4% to 90.0%. Similarly, the Body Model’s F1-score improves from 79.4% to 91.0% on the CI test set and from 74.0% to 89.4% on the PLWA test set. This marked increase in performance confirms that the MH-CSA module is a critical component of the architecture, directly contributing to its high accuracy and robust generalization capabilities as needed in a challenging setting.

**TABLE IV:** Performance comparison of the models with and without the proposed Multi-Head Channel-Spatial Attention (MH-CSA) module. Note: CI: Cognitively Healthy, PLWA: People Living With Alzheimer’s.

| Model | Metric | CI Test Set |  | PLWA Test Set |  |
| --- | --- | --- | --- | --- | --- |
|  |  | Baseline | Proposed | Baseline | Proposed |
| Head | Sensitivity | 67% | 92% | 61% | 90% |
|  | Specificity | 85% | 96% | 81% | 95% |
|  | F1-Score | 70% | 91% | 63% | 90% |
|  | Precision | 73% | 91% | 66% | 90% |
| Body | Sensitivity | 77% | 91% | 71% | 92% |
|  | Specificity | 81% | 95% | 80% | 96% |
|  | F1-Score | 79 % | 91% | 74% | 89% |
|  | Precision | 82% | 91% | 77% | 87% |

## VI. Discussion

The implications of these results for model robustness in minority-class detection scenarios are discussed below with respect to model generalization as required for sensing systems operating under diverse conditions.

The performance metrics shown in Table III, derived from Fig.5, demonstrate strong performance in predicting both head and body poses in this population. When averaged across the three pose prediction classes, all metrics are consistently in the 90%+ region apart from F1 score and precision in the PLWA test group, where this drops to 89% and 87%, respectively. Sensitivity and specificity in both test groups across both head and body pose prediction modes span 91% (sensitivity in the CI body pose) to 96% (specificity in PLWA body pose). F1 score may be argued to be the most insightful metric in a minority class detection framework, which is observed to be within a 2% difference (89-91%) across all test groups and body/head prediction models, again demonstrating a consistent level of performance across both test groups in both head and body pose predictions.

In the head model predictions, it is noticeable that model performance across all metrics is ≈ 1% higher for the CI group compared to the PLWA, although this is unlikely to be of practical significance. This level of metric consistency and differences in model performance across test cohorts is, however, not sustained when comparing both test groups in body model predictions: sensitivity and specificity appear slightly better in the PLWA test group compared to the CI group.

## VII. CONCLUSION

In conclusion, we have presented a novel ML architecture for use in a video sleep monitoring system for determining head and body pose in at-home settings. The model is robust to the presence of occlusion by bed covering on the body and partial occlusion across the head as found in typical sleeping environments. The F1 score for the head(body) model was 91%(91%) for CI test group and 90%(89%) for PLWA. Other test metrics typically fell in the 90%+ range (apart from 87% precision for PLWA). We found that in particular, head pose can only be reliable inferred using a multi-head attention model. Optimal performance for both head and body models was obtained via the use of a Dynamic Cross-Balanced Focal Loss function during training to address class imbalance. Such an approach may be used to accurately assess head and body position in the context of sleep apnea and other sleep phenotypes in which head and body position may be relevant. This model is currently being used in an At-Home video sleep study, and will find wider application in sleep studies both in dedicated sleep study facilities and video studies at-home.

## Data Availability

All data and code used will be made available online after peer-reviewed publication.

## Acknowledgments

The authors thank Damion Lambert, Tegan Ward, Jasmine Ramble, Sara Mahvash Mohamedi and members of the Clinical Research Facility for their help with data collection and curation. They would also like to extend their thanks to the members of Surrey and Borders Partnership (SABP) for help with the recruitment and screening of participants.

